# Global burden of mental health problems among children and adolescents during COVID-19 pandemic: A systematic umbrella review

**DOI:** 10.1101/2022.04.22.22274169

**Authors:** M. Mahbub Hossain, Fazilatun Nesa, Jyoti Das, Roaa Aggad, Samia Tasnim, Mohan Bairwa, Ping Ma, Gilbert Ramirez

**Author notes:** **Corresponding author:** M. Mahbub Hossain, School of Public Health, Texas A&M University, TX 77840.

## Abstract

Mental health problems among children and adolescents are increasingly reported amidst the coronavirus disease (COVID-19) pandemic. In this umbrella review, we aimed to synthesize global evidence on the epidemiologic burden and correlates of child and adolescent mental health (CAMH) problems during this pandemic from existing systematic reviews and meta-analyses. We evaluated 422 citations and identified 17 eligible reviews on the epidemiology of CAMH in the context of COVID-19. Most of the reviews reported a high prevalence of anxiety, depression, sleep disorders, suicidal behavior, stress-related disorders, attention-deficit/hyperactivity disorder, and other mental health problems. Also, factors associated with CAMH such as age, gender, place of residence, educational attainment, household income, sedentary lifestyle, social media and internet use, comorbidities, family relationships, parents’ psychosocial conditions, COVID-19 related experiences, closure of schools, online learning, and social support were reported across reviews. As most studies were cross-sectional and used nonrepresentative samples, future research on representative samples adopting longitudinal and intervention designs is needed. Lastly, multipronged psychosocial care services, policies, and programs are needed to alleviate the burden of CAMH problems during and after this pandemic.

## Introduction

The world has become acquainted with the coronavirus disease 2019 (COVID-19) pandemic that has triggered a wide range of biological, psychological, and socio-economic consequences globally [1–3]. A growing body of empirical research informs a high burden of mental health problems in recent times that is increasingly recognized as a psychiatric epidemic synergistically occurring alongside this pandemic [4, 5]. Several reviews suggested a heavy psychosocial impact of COVID-19 pandemic resulting in mental health problems such as anxiety, depression, posttraumatic stress disorders (PTSD), and sleep disorders [4, 6–9]. An umbrella review of eight systematic reviews and meta-analyses found that preventive measures such as quarantine and isolation for infection prevention may have immediate and prolonged mental health impacts [10]. This evidence suggests ongoing and future mental health crises associated with this pandemic that should be investigated and addressed across various population groups globally.

Mental health problems in childhood and adolescence are unique as they have versatile correlates and may have long-term health biopsychosocial consequences [11–13]. The global prevalence of mental health disorders in children and adolescents has been inconsistent yet high across previous syntheses of research [14, 15]. Specifically, several childhood mental health problems, including depression and anxiety, have become highly prevalent over the past years [8, 11, 14, 15]. For example, a recent study found that the incidence of depression or anxiety in ages 6 to 17 has risen from 5.4% in 2003 to 8.4% in 2011-2012 in the US [16]. Another study that was published in 2010 reported that 1 in 5 US children experience a serious or debilitating mental health problem in their lifetime [17]. Similar studies from Canada [18, 19], Europe [20, 21], and Australia [22, 23] suggest comparative high burden of child and adolescent mental health (CAMH) problems. Moreover, countries with limited resources, mostly those in the global south, often lack high-quality evidence on mental health epidemiology. Yet, emerging literature from many low- and middle-income countries suggests a high burden of CAMH problems in those contexts that is a global mental health concern [24–26]. These numbers are expected to increase amidst and after the COVID-19 pandemic.

The impacts of the COVID-19 pandemic on the mental health outcomes of children have drawn considerable attention amongst global researchers. Children and adolescents constitute the most vulnerable population affected by the preventive measures for COVID-19, such as the closure of schools, leading to reduced interaction with peers, and lessened opportunities for physical activity and exploration [27, 28]. These effects are expected to have adverse impacts on the welfare and mental health of children through sleeping disorders, depression, stress and anxiety [5, 28]. Studies conducted across the globe provide evidence of the detrimental impact of the pandemic on the mental health of adolescents [29]. During the COVID-19 pandemic lockdown, the percentage of Bangladeshi children who suffered from mental health disturbances ranged from 43% for subthreshold disturbances to 7.2% for severe disturbances [30]. In Turkey, children of health workers had significantly higher anxiety levels measured using the State-Trait Anxiety Inventory for Children (STAI-C) than non-health workers’ children [31]. In Ireland, a qualitative study showed that adolescents and children experienced increased rates of social isolation, maladaptive behavior, anxiety, and depression during the pandemic [32]. Moreover, the pandemic resulted in a significant psychosocial impact on Chinese adolescents and children in terms of emotional distress, with a prevalence of 24.9% for anxiety, 19.7% for depression, and 15.2% for stress [33]. Another systematic review of Chinese studies showed that the prevalence of COVID-19 related posttraumatic stress, sleep disorders, anxiety, and depression were 48%, 44%, 26%, and 29%, respectively [34].

An increasing body of primary studies has facilitated the development of many such reviews that should be synthesized to provide an extensive overview of all CAMH problems and their associated factors. Such systematic overview of reviews, also known as umbrella reviews, have become increasingly useful in understanding the evidence landscape in a given domain [35]. The findings of an umbrella review can offer a “bird’s eye” view of a scientific field that may help evidence-based decision-making, synthesizing future evidence, and conducting primary research on least-explored areas of knowledge and practice [35–37]. This umbrella review aimed to summarize the global evidence on the epidemiological burden and correlates of mental health problems among children and adolescents during the COVID-19 pandemic from existing systematic reviews and meta-analyses. The results of this review can help understand and address mental health problems among children and adolescents in this pandemic and future public health emergencies.

## Methodology

### Guidelines, data sources, and search strategy

In this umbrella review, we have followed the Preferred Reporting Items for Systematic Reviews and Meta-analyses (PRISMA) guideline [38] and the Joanna Briggs Institute (JBI) recommendations [39]. The conduct of an umbrella review aims to address the broader scope of a scholarly topic and related issues presenting a wide picture of existing evidence. Umbrella reviews identify and examine evidence from existing systematically conducted reviews through a rigorous process. In this umbrella review, we conducted an extensive literature search in PubMed/MEDLINE, APA PsycInfo, Academic Search Ultimate, Cumulative Index to Nursing and Allied Health Literature (CINAHL), Health Source-Nursing/Academic Edition, Health Policy Reference Center, and Web of Science using a set of keywords as outlined in Table 1.

**Table 1:** Keywords and search strategy.

| **Search query** | **Search topic** | **Search keywords (titles, abstracts, general keywords, and subject headings)** |
| --- | --- | --- |
| **1** | Condition/<br>outcome of<br>interest | “mental health” OR “mental wellbeing” OR “mental well-being”<br>OR “mental disorder*” OR “mental illness” OR “psychiatr*” OR<br>“psychological health” OR “psychological distress” OR<br>“psychological impact” OR “psychological outcomes” OR<br>“psychological consequence*” OR “psychological comorbid*” OR<br>“psychosocial problem*” OR “behavioral problem*” OR<br>“behavioral disorder*” OR “cognitive disorder*” OR “cognitive<br>impairment*” OR “emotional distress” OR “depression” OR<br>“depressive disorder*” OR “anxiety” OR “PTSD” OR “PTSS” OR<br>“posttraumatic” OR “posttraumatic” OR “addiction” OR<br>“substance use disorders” OR “mood disorder*” OR “affective<br>disorder*” OR “DSM*” OR “psychosis” OR “psychotic” OR<br>“oppositional defiant disorder” OR “hyperactiv*” OR “conduct<br>disorder” OR “obsess*” OR “phobi*” OR “schizophren*” OR<br>“bipolar disorder” OR “anorexia” OR “bulimi*” OR “challenging<br>behav*” |
| **2** | Population | “child*” OR “adolescen*” OR “young people” OR “school<br>students” OR “youth” OR “teen*” |
| **3** | Exposure/<br>Context | “Coronavirus” OR “COVID-19” OR “COVID19” OR “SARS-CoV-2”<br>OR “2019 novel coronavirus” OR “2019-nCoV” |
| **4** | Phenomenon | “prevalence” OR “incidence” OR “rate” OR “diagno*” OR “disease<br>burden” OR “epidemiolog*” OR “frequen*” OR “determinant*” OR<br>“predictor*” OR “correlate*” OR “risk factors” OR “protective<br>factors” OR “associated factors” |
| **5** | Study types | “systematic review” OR “systematic literature review” OR “meta-<br>analy*” OR “metaanaly*” OR “quantitative review” OR<br>“qualitative review” OR “meta-synthesis” OR “pooled prevalence”<br>OR “pooled estimate*” OR “scoping review” OR “critical review”<br>OR “integrative review” OR “evidence-based review” OR<br>“synthesized evidence” |
| **Final**<br>**search**<br>**query** | Intersection<br>of four topics | 1 AND 2 AND 3 AND 4 AND 5 |

### Inclusion criteria

#### Participants

This review focused on children and adolescents irrespective of the age range adopted in systematic reviews and the primary studies included in the same. For studies with mixed participants such as adolescents with young adults or children as a part of other population groups, a decision was made prior to conducting the review about including a review if at least 70% of its studies focused on children or adolescents.

#### Phenomenon of interest

This review studied a wide range of mental health problems, including but not limited to psychiatric disorders enlisted in the Diagnostic and Statistical Manual of Mental Disorders (DSM–5) and the International Classification of Diseases (ICD-10) Classification of Mental and Behavioral Disorders. To make the evidence inclusive of any problem that affects mental health and wellbeing, this review considered research that emphasized psychosocial problems alongside psychiatric conditions.

#### Context

The context of this review was the COVID-19 pandemic. Therefore, studies conducted during and in the context of this pandemic were considered eligible for this review. If a review used data collected beyond the scope of this pandemic, it was considered ineligible for inclusion. Moreover, studies with mixed data from COVID-19 and previous infectious outbreaks were reviewed carefully. In such cases, a study was considered eligible if at least 70% of the respondents on included studies informed evidence in the context of COVID-19.

#### Types of studies

Any review article with a systematic approach to retrieve and synthesize evidence was considered eligible for inclusion in this umbrella review. This decision was made considering methodological differences within systematically conducted review articles. Therefore, systematic literature reviews, scoping reviews, and meta-analyses were included, whereas primary studies or narrative reviews without a clear review method were excluded. Moreover, articles were included in the full text was available in the English language. Lastly, studies published from January 1, 2020, to December 31, 2021, were primarily included in this review, and the search was last updated on March 12, 2022.

### Study selection

We collated and uploaded all identified citations were to the cloud-based screening tool Rayyan. Two reviewers independently reviewed those citations using the above-mentioned criteria. All conflicts were addressed through a discussion, and the opinion of a third reviewer was taken whenever it was necessary. Articles that appeared to be qualified at this stage were considered for full-text review, and a final set of articles meeting all eligibility criteria was retained for data extraction and synthesis.

### Data extraction and synthesis

We used a standardized data extraction form that included key variables of interest such as the bibliographic information of each article, the type of the reviews, number of primary studies, databases accessed in respective reviews, characteristics of the study samples and populations, epidemiologic measures used in those studies, and finally, the burden of mental health problems and factors associated with the same. Two reviewers (FN and AD) conducted the data extraction independently in the supervision of a third reviewer (ST). The extracted data were further reviewed for accuracy and quality by another reviewer (MMH). As this review included a wide range of systematically conducted reviews with and without quantitative synthesis, there was marked heterogeneity in the methods and outcomes of the included reviews. For this reason, a narrative synthesis of overall evidence on the burden of and factors associated with mental health problems in children and adolescents during the COVID-19 pandemic was presented in this review.

### Assessment of methodological quality

In this umbrella review, we used the JBI checklist for the critical appraisal of systematic reviews and research syntheses to assess the methodological quality of studies included in this umbrella review [35, 39]. This checklist has ten items evaluating different methodological aspects of a systematically conducted review, including the appropriateness of the search strategies, the approach to synthesizing evidence, potentials sources of biases, and prospects for future research and policymaking. We used this ten-items checklist and allocated one point for each item. So, the overall score of a study can range from zero to ten. In this review, studies receiving zero to four, five to seven, and eight to ten points were categorized as the low, medium, and high-quality studies, respectively.

## Results

### Overview of the included studies

We found a total of 422 citations using the search strategy described earlier (See Figure 1 for the review process). After eliminating 181 duplicate citations, we retained 241 unique citations and reviewed their titles and abstracts. A total of 24 citations met our preliminary criteria and were retained for full-text review. After full-text evaluation, 17 systematic reviews [29, 34, 48–54, 40–47], including 6 meta-analytic reviews [34, 42, 43, 46, 49, 51] met all criteria of this umbrella review and were selected for further analysis (see Table 2).

**Figure 1:**
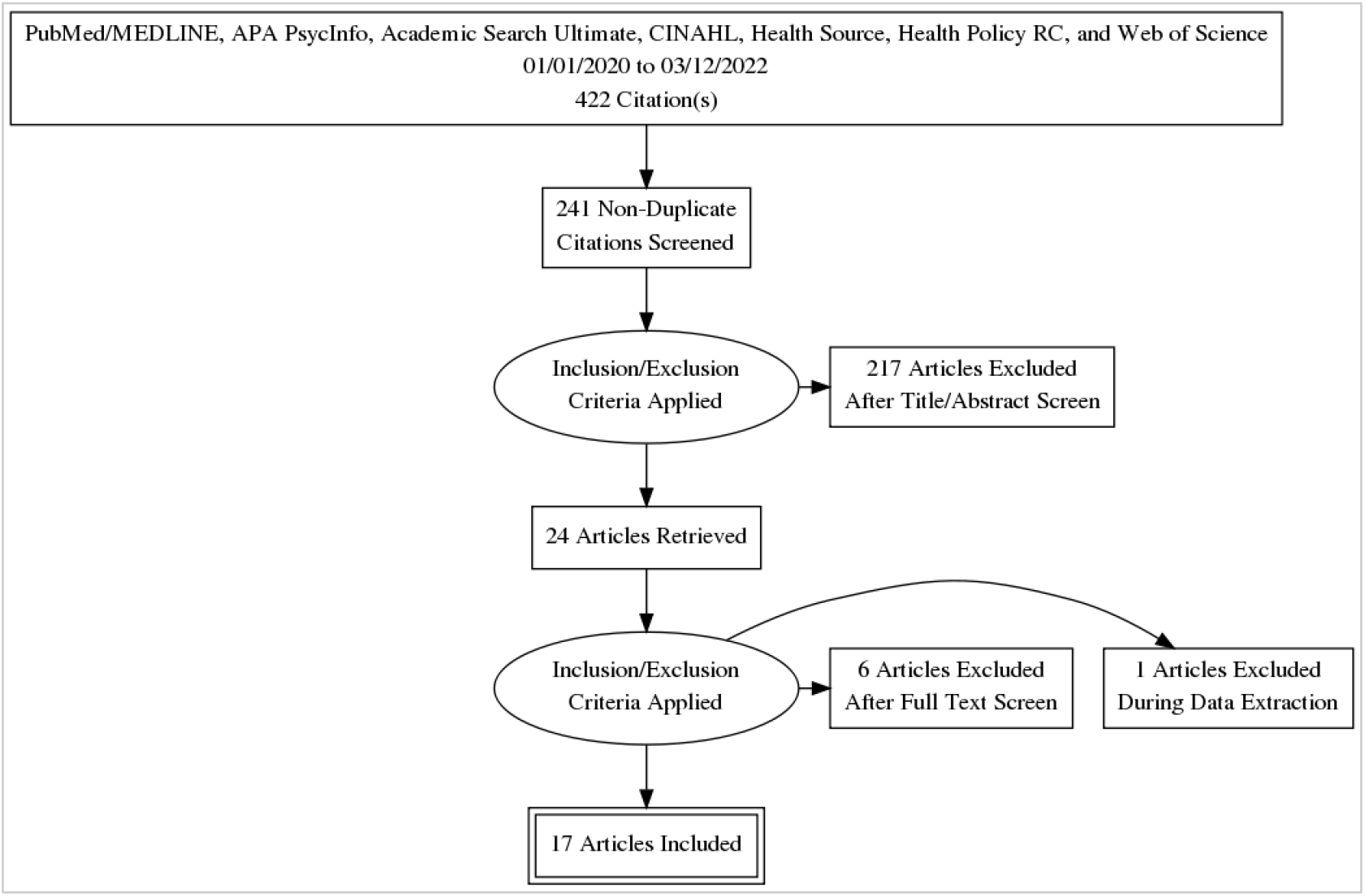
Flow diagram of the umbrella review process.

**Table 2:**
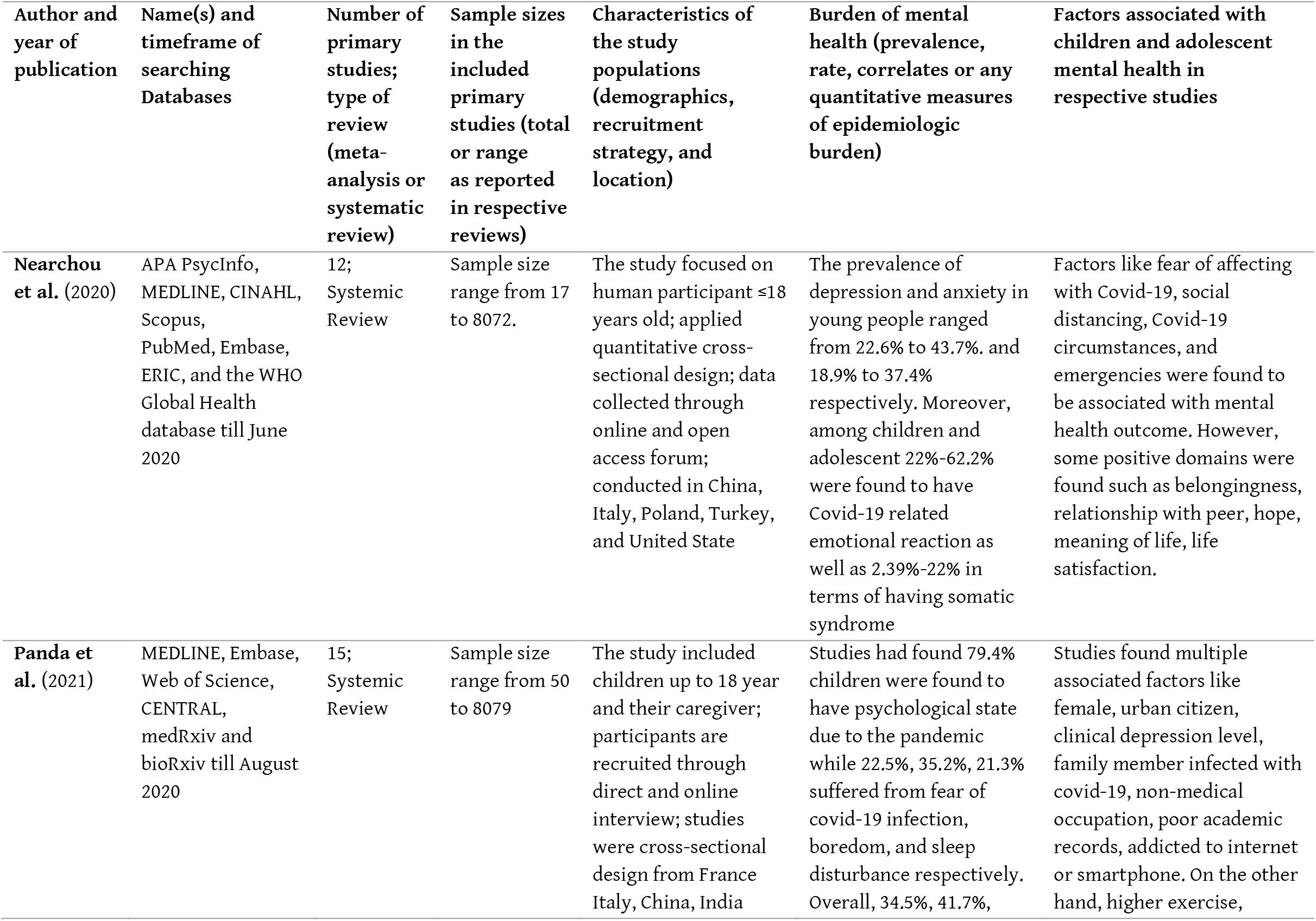
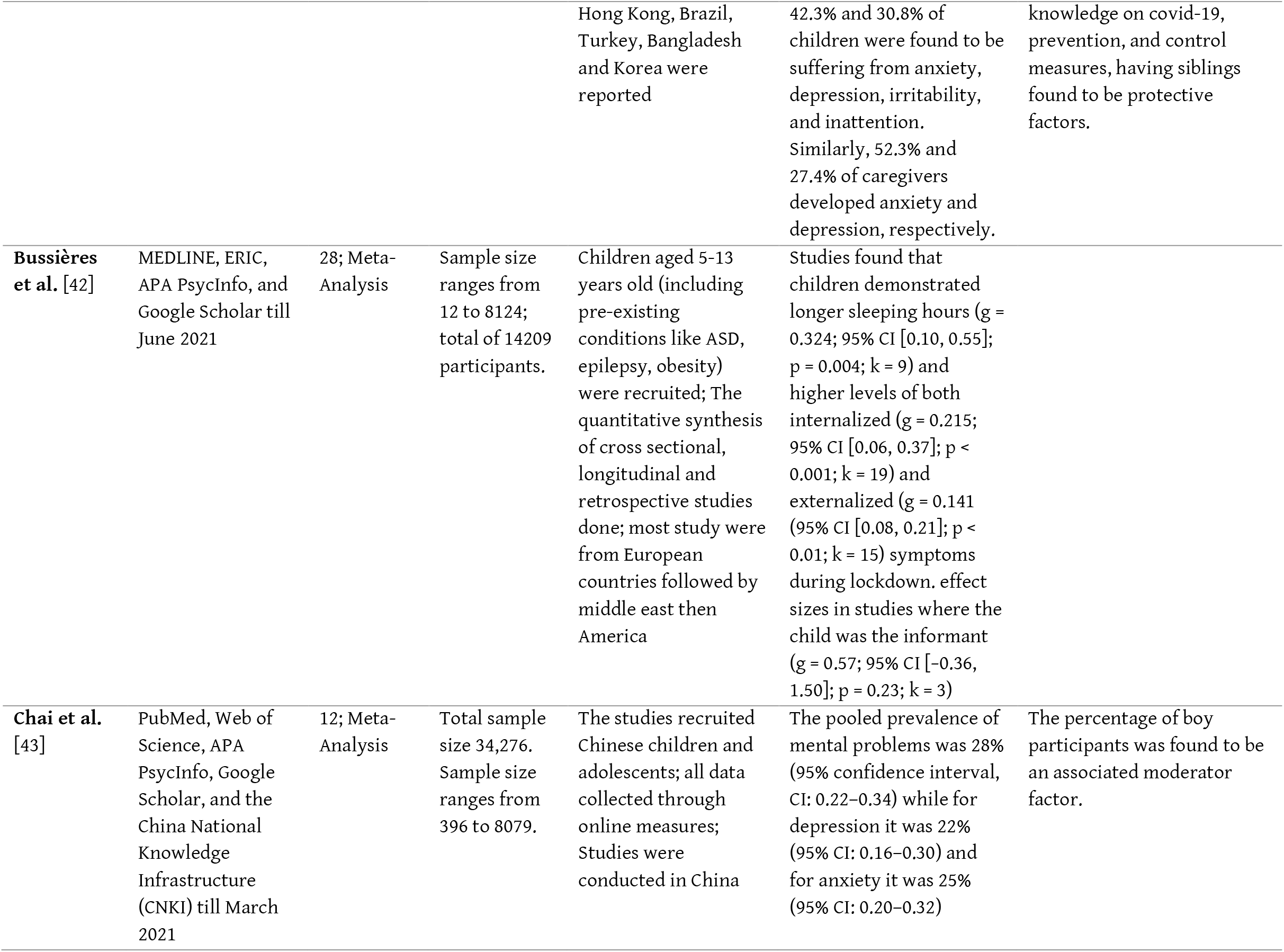
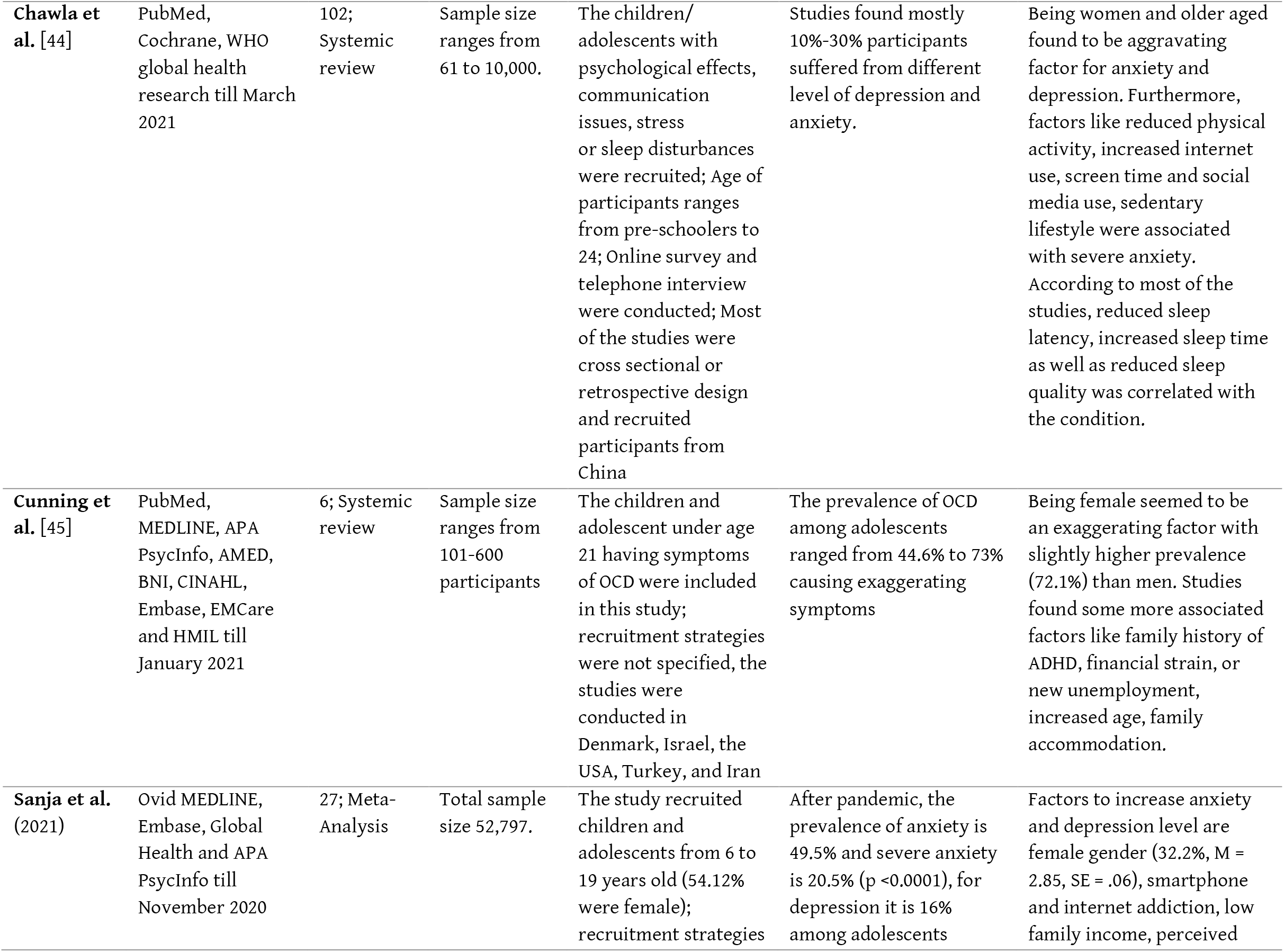
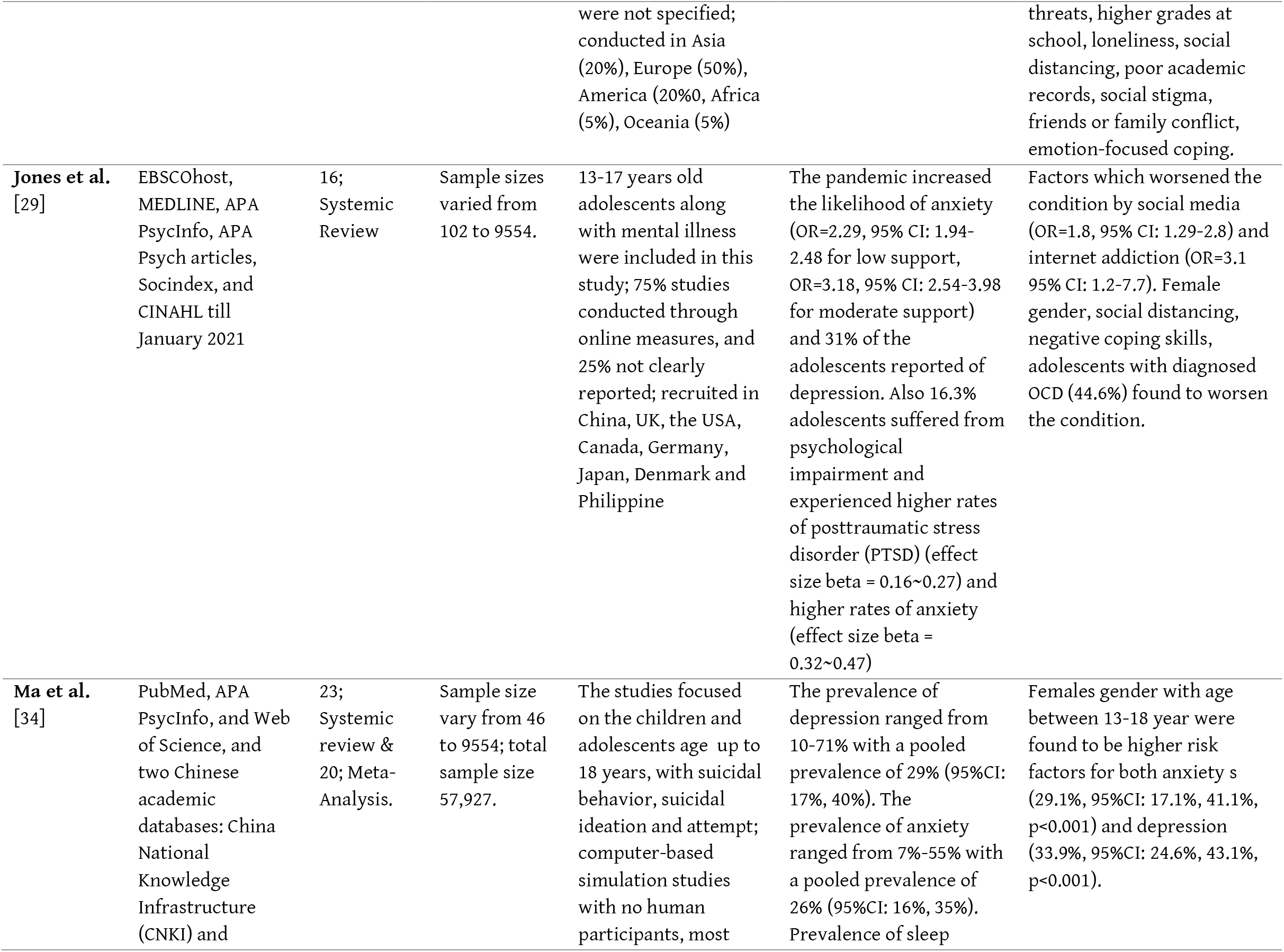
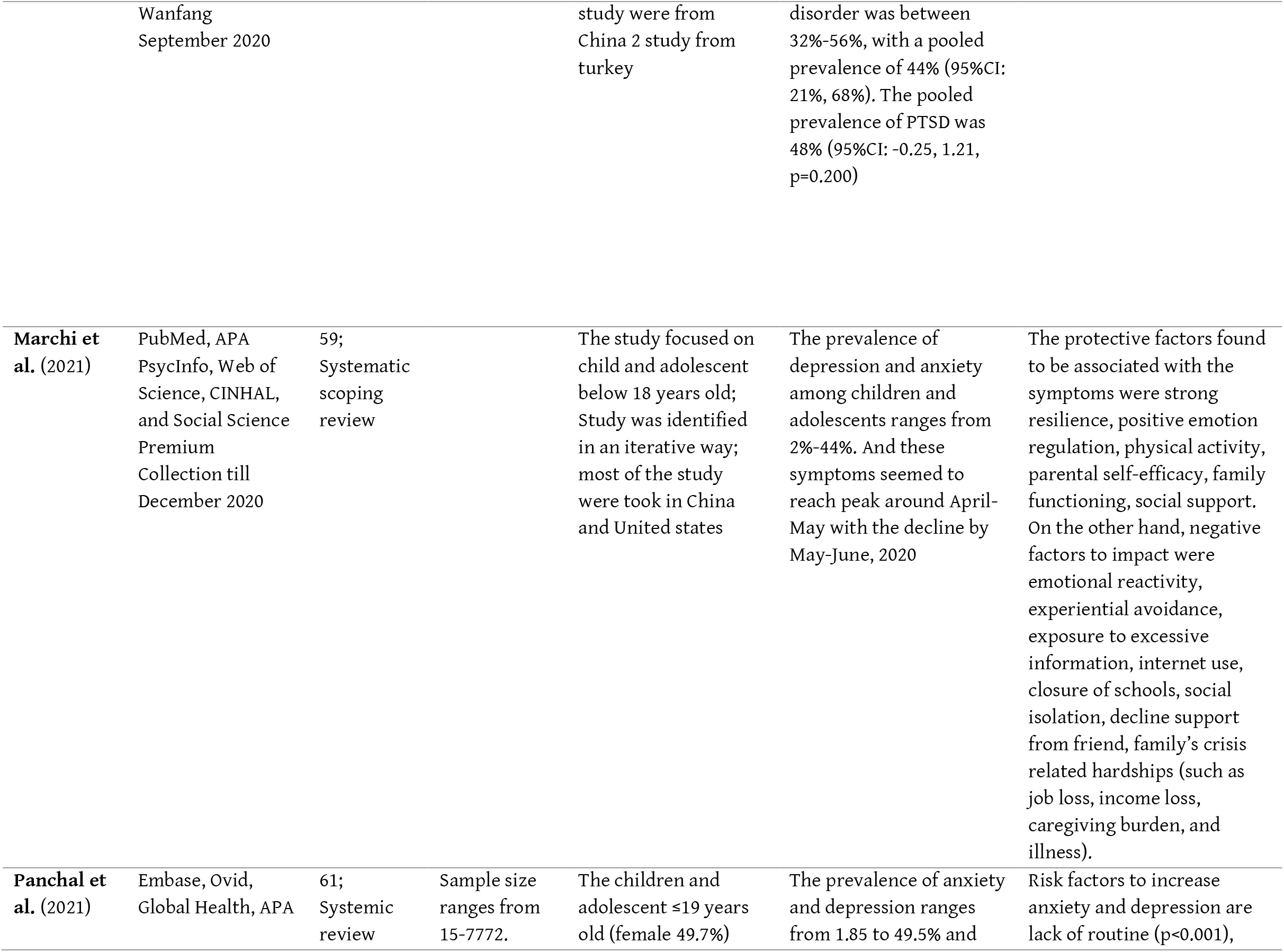
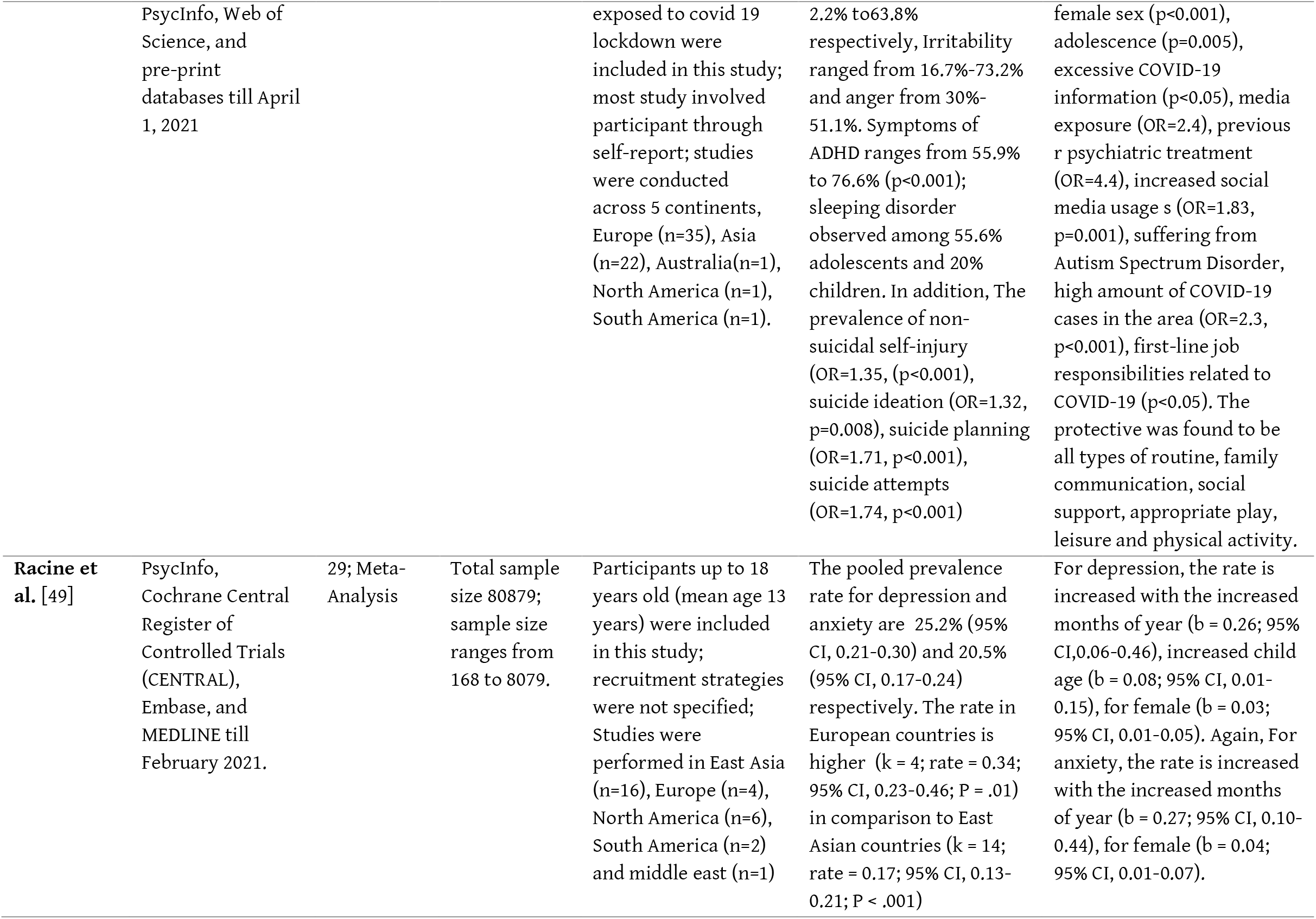
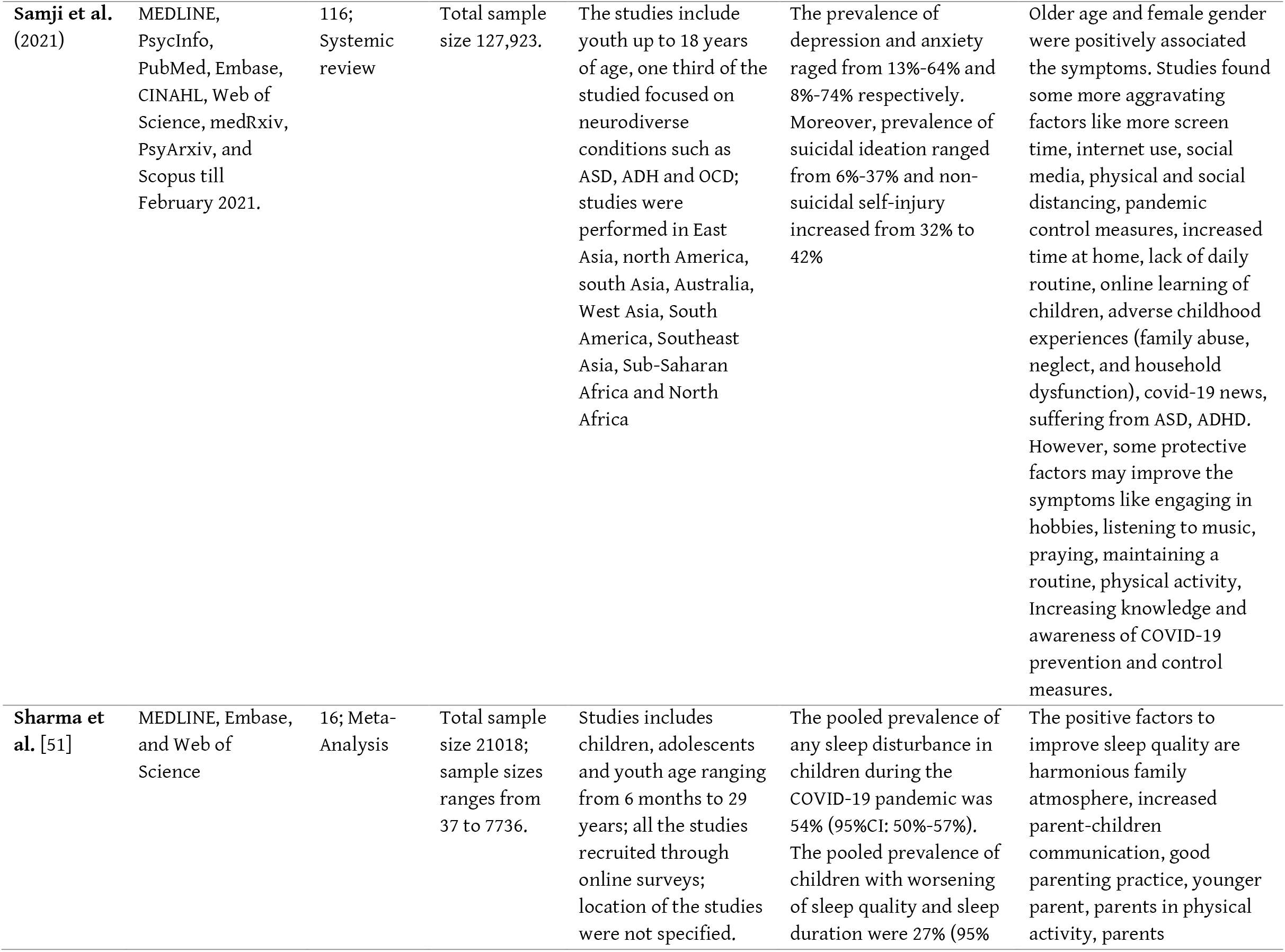
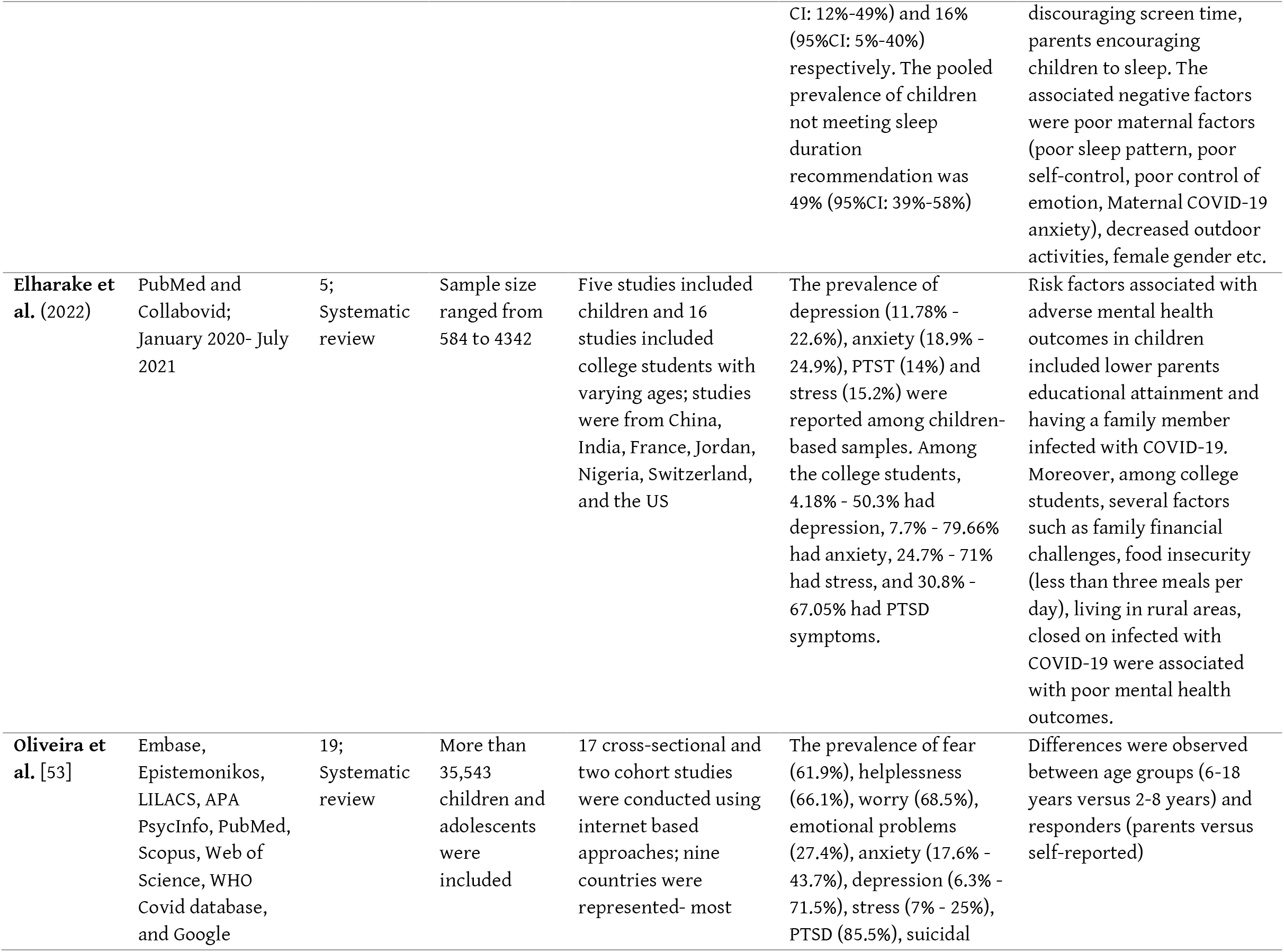
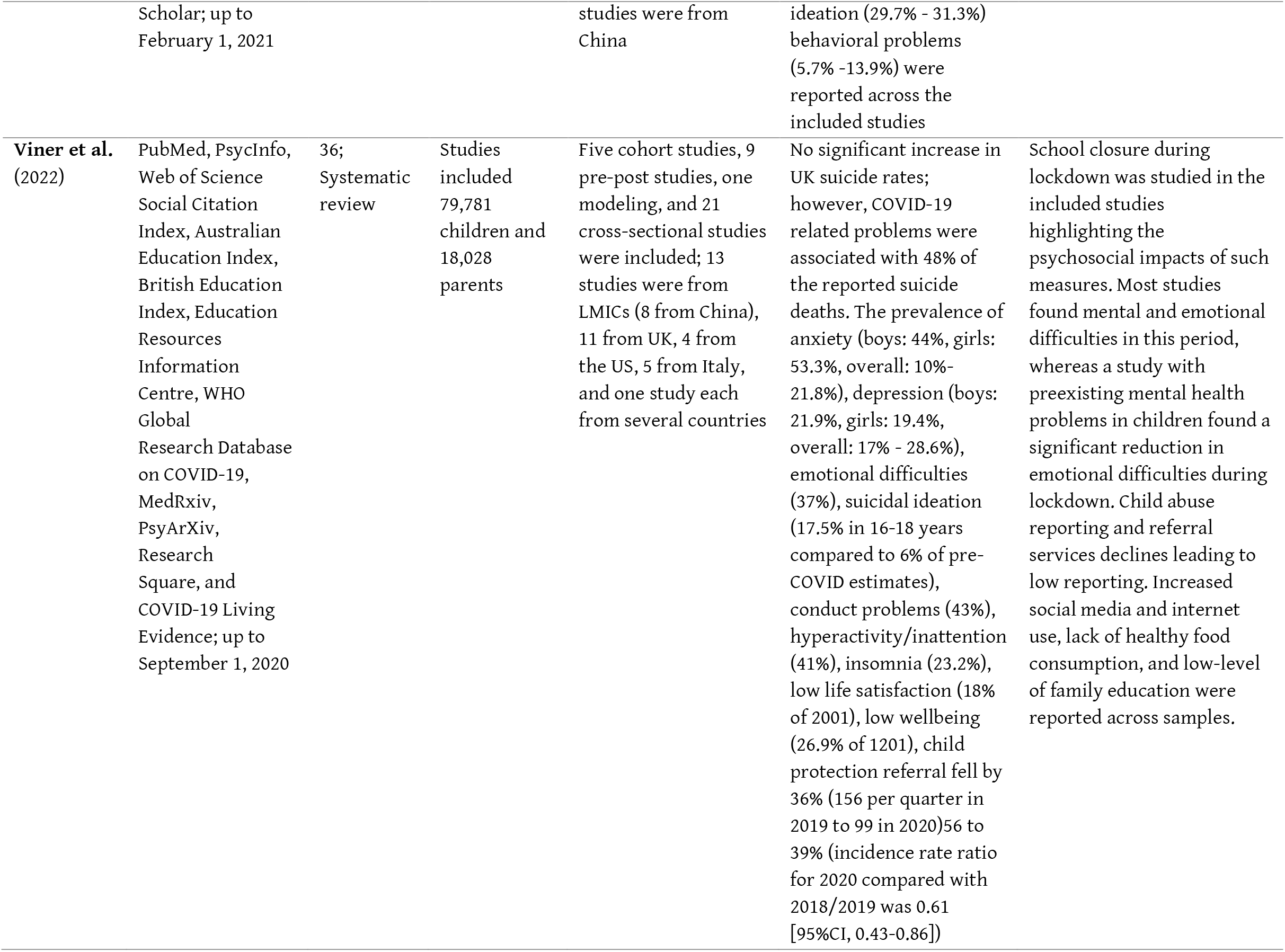
Overview of evidence from existing systematic reviews and meta-analyses on CAMH problems during COVID-19.

### Characteristics of the included studies

Seventeen reviews included in this umbrella review used different databases for retrieving primary studies. The number of databases ranged from 2 to 11 with a median number of 5 databases, whereas MEDLINE and WHO COVID-19 databases are the leading sources of information across multiple reviews.

The number of primary studies included in different reviews ranged from 5 to 102. Moreover, the sample size across studies ranged from 12 to 1,199,320 in systematic reviews, whereas a meta-analytic review used a pooled sample of 80,879 participants [49]. The majority of the primary studies were cross-sectional, and online data collection measures were widely used across studies. China was the most common source of primary studies, followed by the North American and European countries. Limited primary studies were reported from LMICs such as India, Brazil, Bangladesh, and countries from Sub-Saharan Africa [41, 46, 48–50, 52].

The methodological quality of the reviews was assessed using the JBI critical appraisal checklist (See Supplementary Material). Most of the reviews (n = 13) included in this umbrella review were of high quality [29, 34, 51, 53, 54, 40–44, 48–50], whereas the remaining (n = 4) reviews had medium quality [45–47, 52]. None of the reviews were identified to have low methodological quality as per the JBI checklist.

### Epidemiological burden of CAMH during COVID-19

#### Anxiety

Thirteen reviews reported a varying burden of anxiety among children and adolescents during this pandemic [29, 34, 51, 53, 54, 40, 43, 44, 46–50]. The prevalence of anxiety ranged from 1.85% to 74% across primary studies included in the reviews. For example, studies reported in the review by Panchal et al. (2021) found the prevalence of anxiety ranging from 1.85% to 49.5%. Another review by Samji et al. (2021) reported the prevalence of anxiety between 8% and 74%. A meta-analytic review by Racine et al. (2021) found that 20.5% (95% CI: 17% - 24%) participants had anxiety disorders during this pandemic.

#### Depression

Among the included studies, thirteen reviews and meta-analyses evaluated a high prevalence of depression among children and adolescents ranging from 2% to 71.5% [29, 34, 52–54, 41, 43, 44, 46–50]. For example, Marchi et al. (2021) found that the prevalence of depression was between 2% and 4% across the reviewed literature. Another review by Oliveira et al. (2022) reported the prevalence of depression between 6.3% and 71.5%. A meta-analytic review by Ma et al. (2021) reported the pooled prevalence of depression as 29% (95% CI: 17% - 40%) among children and adolescents.

#### Sleep disorders

Seven reviews reported varying magnitude of sleep disorders among children and adolescents during this pandemic [34, 41, 42, 44, 48, 51, 54]. For example, Panchal et al. (2021) reported that the prevalence of sleep disorders was 20% and 55.9% among children and adolescents, respectively. Another review by Sharma et al. (2021) reported a pooled prevalence of any sleep disorders as 54% (95% CI: 50% - 57%), whereas the pooled prevalence of worsening sleep quality was 27% (95% CI: 12% - 49%) and the pooled estimate of abnormal sleep duration was 16% (95% CI: 5% - 40%).

#### Trauma and stress-related disorders

Four reviews examined the burden of posttraumatic stress disorders in different samples of children and adolescents [29, 34, 52, 53]. For example, Ma et al. [34] found a pooled prevalence of PTSD as 48%, whereas another review by Jones et al. [29] found that adolescents experienced a high effect size of PTSD (beta = 0.16 to 0.27). Another review by Elharake et al. [52] found that 30.8% to 67.05% of the study samples had PTSD symptoms among young people amidst the COVID-19 pandemic. Moreover, three reviews have reported a varying burden of psychological distress and stress-related disorders [41, 52, 53]. For example, Oliveira et al. (2022) reported that 7% to 25% of children and adolescents experienced stress-related disorders amidst this pandemic.

#### Suicidal thoughts, ideations, and behavior

Four reviews studied the burden of suicidal thoughts, ideations, and self-harm behavior among children and adolescents during this pandemic [48, 50, 53, 54]. For example, Panchal et al. (2021) reported a high prevalence of suicide ideation (OR = 1.32, p = 0.008), suicide planning (OR = 1.71, p < 0.001), suicide attempts (OR = 1.74, p < 0.001), and non-suicidal self-harm (OR = 1.35, p < 0.001). Another review by Samji et al. (2021) found that the prevalence of suicidal ideation ranged from 6% to 37%, and non-suicidal self-increased from 32% to 42% during this pandemic. Viner et al. (2022) found that the prevalence of suicidal ideation increased to 17.5% among adolescents aged 16-18 years compared to 6% of pre-COVID estimates.

#### Attention-deficit/hyperactivity disorder

Several empirical studies reported the burden of attention-deficit/hyperactivity disorder in children and adolescents during this pandemic that was synthesized in three reviews [41, 48, 54]. Viner et al. (2022) found that 41% of the participating children had hyperactivity/inattention, whereas Panchal et al. (2021) reported that the symptoms of ADHD ranged from 55.9% to 76.6% (p < 0.001) among children and adolescents across multiple studies. Panda et al. (2021) found that 30.8% of the participating children had attention-deficit symptoms alongside other mental health problems.

#### Other mental health problems

Several other mental health problems were reported in multiple reviews. Cunning et al. [45] reported that the prevalence of OCD among adolescents ranged from 44.6% to 73% causing exaggerating symptoms. Moreover, emotional disorders were reported by three reviews that found 22% to 62.2% of children and adolescents experienced emotional difficulties during this pandemic [40, 53, 54]. A review by Nearchou et al. (2020) found that 2.39% to 37% of participants had somatic syndrome. Furthermore, psychological problems such as fear of COVID-19 [41, 53], boredom [41], irritability [41, 48], internalized and externalized problems [42], helplessness [53], conduct problems [54], and low life satisfaction [54] were reported across reviews. In addition, child protection referral fell by 36% (156 per quarter in 2019 to 99 in 2020) to 39% (incidence rate ratio for 2020 compared with 2018/2019 was 0.61 [95%CI, 0.43-0.86]), suggesting a potential burden of child abuse and related mental health problems [54]. Overall mental and psychosocial health problems deteriorated across all reviews that included children, adolescents, and their family caregivers in respective empirical studies.

### Correlates of mental health problems in children and adolescents during COVID-19

Multiple factors associated with mental health problems were identified across multiple reviews that affected the mental health of children and adolescents during this pandemic. The age of the children and adolescents was reported to be associated with mental health problems in different studies. For example, Chawla et al. (2021) found that older children and adolescents was an aggravating factor for anxiety and depression. Increased age was also reported as a risk factor by Cunning and Hodes [45], Samji et al. [50], and Ma et al. (2021). Specifically, Racine et al. [49] found that increased child age was associated with a higher burden of depression (b = 0.08, 95% CI: 0.01 – 0.15). Moreover, differences in mental health outcomes were observed between participants aged 2-8 years and those aged 6-18 years in a review by Oliveira et al. (2022).

Gender was associated with mental health outcomes as reported in several reviews. Chai et al. reported the percentage of boys was a moderator of mental health outcomes across study samples. In contrast, several reviews reported girls had a higher burden of mental health problems during this pandemic. For example, female gender was associated with increased anxiety and depression level (32.2%, M = 2.85, SE = .06) as reported by Sanja et al. [46]. Another review by Panchal et al. [48] found that the female gender was significantly associated with anxiety and depression (p < .001). Racine et al. [49] reported that female participants had higher rates of depression (b = 0.03, 95% CI: 0.01 – 0.05).

Urban residence was found to be associated with mental health outcomes in a review by Panda et al. [41], whereas Elharake et al. [52] reported poor mental health outcomes in college students living in rural areas. Moreover, the time of the study appeared to affect the study outcomes. For example, Racine et al. [49] found that later months of the year was associated with higher burden of anxiety (b = 0.27, 95% CI: 0.10 – 0.44) and depression (b = 0.26, 95% CI: 0.06 – 0.46). Furthermore, poor academic records were associated with adverse mental health outcomes in one review [41], whereas another review found that higher academic grades were associated with anxiety and depression [46].

Several reviews synthesized evidence on behavioral and lifestyle-related factors associated with mental health problems. For example, Chawla et al. [44] found that a sedentary lifestyle, increased screen time, and lack of exercise were associated with depression and anxiety. Physical activity and regular exercise were associated with improved mental health across multiple reviews [41, 44, 47, 48, 50, 51]. Staying home for increased duration and lack of daily routine affected mental health in many children and adolescents, as reported by Panchal et al. (2021) and Samji et al. [50]. A Review by Viner et al. [54] found that a lack of healthy food consumption was associated with adverse mental health outcomes among children amidst this pandemic. Moreover, better emotional regulation and psychological resilience were associated with improved mental health outcomes. Furthermore, better knowledge about the pandemic was associated with better mental health among children and adolescents [47], whereas excessive information was negatively associated with mental health, as reported by Panchal et al. (2021).

Individual behaviors such as hobbies, praying, and listening to music were reported to be associated with positive mental health [50]. Internet and smartphone addiction appeared to be a common risk factor associated with mental health problems reported in several reviews [29, 41, 44, 46–48, 50, 51, 54]. Moreover, the past history of mental health problems was associated with adverse mental health outcomes, as reported in several reviews [29, 45, 48, 50, 54]. Severe events related to COVID-19 affected mental health among many children and adolescents. For example, Panda et al. (2021) found that having a family member who was infected with COVID-19 was associated with adverse psychological outcomes. Another review by Panchal et al. [48] reported an increased number of COVID-19 cases within the same geographic area affected mental health (OR = 2.3, p < 0.001) in children and adolescents.

Several family issues were reported to be associated with mental health outcomes during this pandemic. Nearchou et al. [40] found that belongingness and better relationships with closed ones were associated with improved mental health. Parental self-efficacy and family functioning were protective factors for mental health, as reported by Marchi et al. [47]. Moreover, adverse childhood and family experiences such as household dysfunction, family neglect, and child abuse were associated with poor mental health outcomes [50]. Moreover, the parents education and responsiveness were associated with the mental health of children and adolescents [51–54]. In addition, family financial crises affected the mental health of children and adolescents in many reviews. For example, unemployment and financial constraints affected families and the mental health of young people [45]. Elharake et al. [52] found that family financial challenges and food insecurity (less than three meals per day) affected mental health in children and college students.

School closure was associated with a wide range of mental health outcomes, as studied in a review by Viner et al. [54] and Marchi et al. [47]. Specifically, Viner et al. [54] found that child abuse referral reports declined during this pandemic due to school closure suggesting interrupted services in those contexts. As many institutions transitioned to online learning systems, Samji et al. [50] found such online learning to be associated with adverse mental health outcomes in children and adolescents.

Social factors such as social isolation and distancing, social stigma, and social support were associated with overall mental health outcomes in many reviews [29, 46–48, 50]. For example, social distancing was associated with adverse mental health outcomes in a review by Jones et al. [29]. Moreover, Samji et al. [50] reported that social distancing was associated with poor mental health outcomes. Both social stigma and social distancing were related to a higher burden of anxiety in a review by Sanja et al. Furthermore, Panchal et al. [48] and Marchi et al. [47] found social support to have a protective role on mental health outcomes in children and adolescents.

## Discussion

This umbrella review provides a systematic overview of global evidence on CAMH during the COVID-19 pandemic from existing systematic reviews and meta-analyses. To the best of our knowledge, this is one of the few umbrella reviews on mental health in COVID-19 [55, 56] and the first one emphasizing CAMH. The findings of this review may facilitate mental and child health policymaking and practice and inform further research strengthening the global knowledge base. In this review, we found a varying burden of several mental health problems such as anxiety, depression, sleep disorders, PTSD, suicidal behavior, psychological distress, and other psychosocial challenges among children and adolescents. Moreover, we identified several critical factors associated with mental health problems in this young population. Individual social demographic factors associated with mental health outcomes included older age of the children and adolescents, female gender, location of the residence, educational attainment, sedentary lifestyle, lack of routine activities, spending more time at home, social media addiction, Internet overuse, knowledge about pandemic or excessive information, emotional regulation, resilience, self-efficacy, physical activity and exercise behavior, recreational activities, spending time on hobbies, and history of mental health problems. Furthermore, several family-related factors associated with mental health were identified, which included belongingness and relationship with family members, parents’ education, household income and economic hardship, adverse childhood and family experience, and challenging family relationships. In addition, community and social factors related to mental health included social distancing practices, social support, social stigma, closure of the schools, online learning activities, and COVID-19 related life events and preventive measures. These factors influenced mental health outcomes among children and adolescents in different contexts as identified in existing reviews and respective primary studies included within those reviews.

Most studies reported a high prevalence of CAMH problems across contexts suggesting a heavy psychosocial impact of this pandemic that is consistent with other reviews conducted in different populations [5, 55–58]. Socioecological perspectives may inform the complex epidemiology of these crises emerging at different levels. Children and adolescents experienced an unusual interruption in their everyday lives that included prolonged stay within home, altered relationship dynamics with their caregivers and family members, less-frequent interactions with peers and relatives, lack of access to outdoor activities, changed nutritional and lifestyle behaviors and other changes at the household level [59, 60]. These problems compounded the effects of community and social changes associated with this pandemic, which included social distancing, closure of schools and institutions, and interruption of health and social services [54, 61, 62]. Lastly, universal challenges such as socioeconomic problems and health hazards of COVID-19 impacted children, adolescents, and their families that also affected mental health and wellbeing [5, 47, 63]. Taken together, a variety of psychosocial stressors emerged during this pandemic, whereas mental health promotion and protective services were mostly disrupted or unavailable. These factors affected biopsychosocial health and wellbeing across populations resulting in a high burden of CAMH problems that require clinical, public health, social, and policy considerations.

The overall evidence on the epidemiologic burden of CAMH and associated factors reflected high heterogeneity that can be attributed to several factors. First, earlier studies on psychosocial impacts were conducted in China [34, 45], whereas a majority of overall studies were from the US, UK, Canada, and several high-income countries [42, 49, 52, 54]. Most reviews informed a scarcity of evidence from the global south reflecting a critical knowledge gap in those contexts. Moreover, online measures of collecting data and non-standardized scales were widely used in cross-sectional studies reported across reviews [40, 44, 45, 47, 52], which may not provide representative estimates on CAMH impacts of COVID-19. Therefore, standardized measurements approaches should be used in representative samples and preferably in longitudinal and interventions studies that more accurately reflect the psychosocial impacts locally and globally. A brief overview of the recommendations for future research on CAMH is provided in Table 3-A.

**Table 3:**
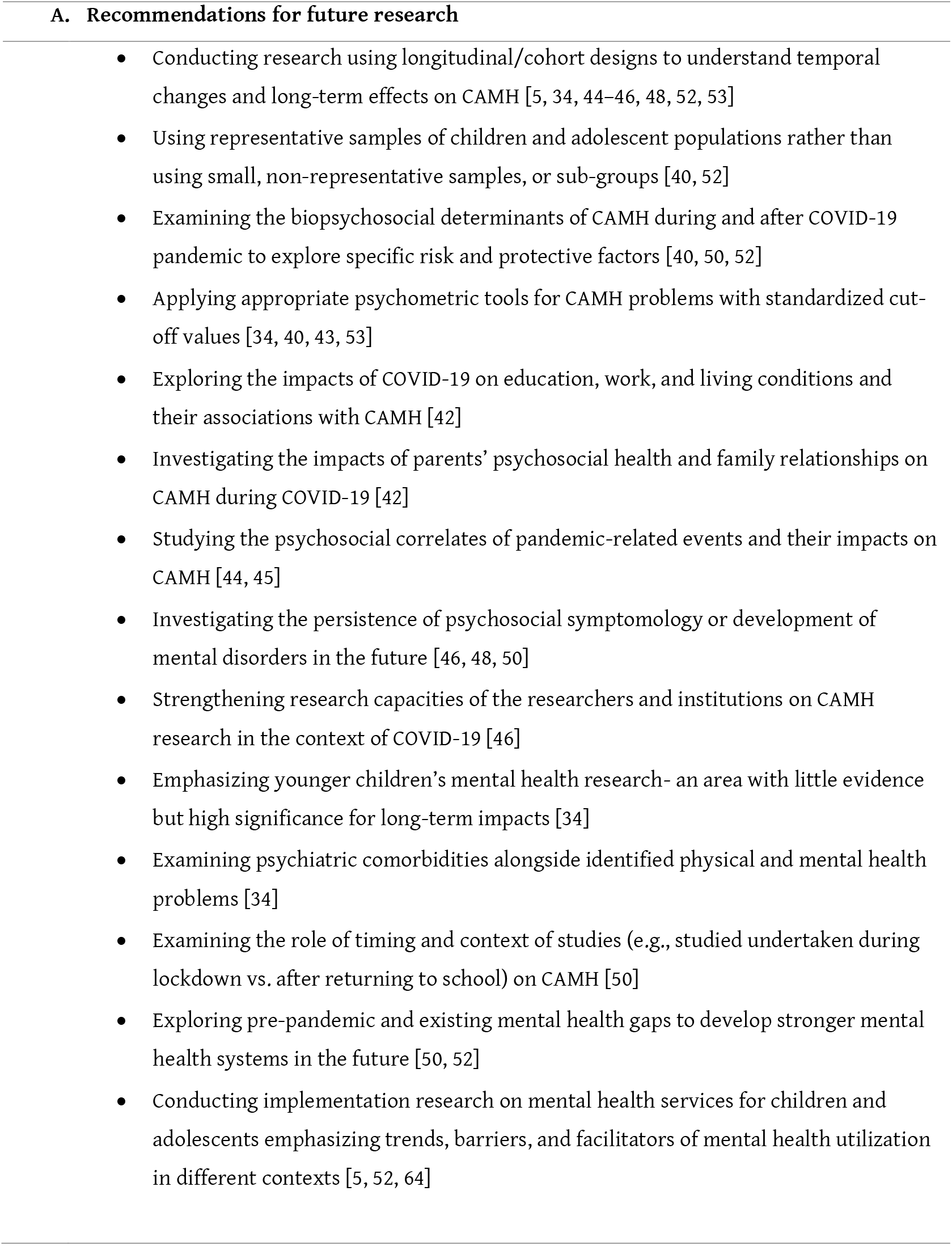
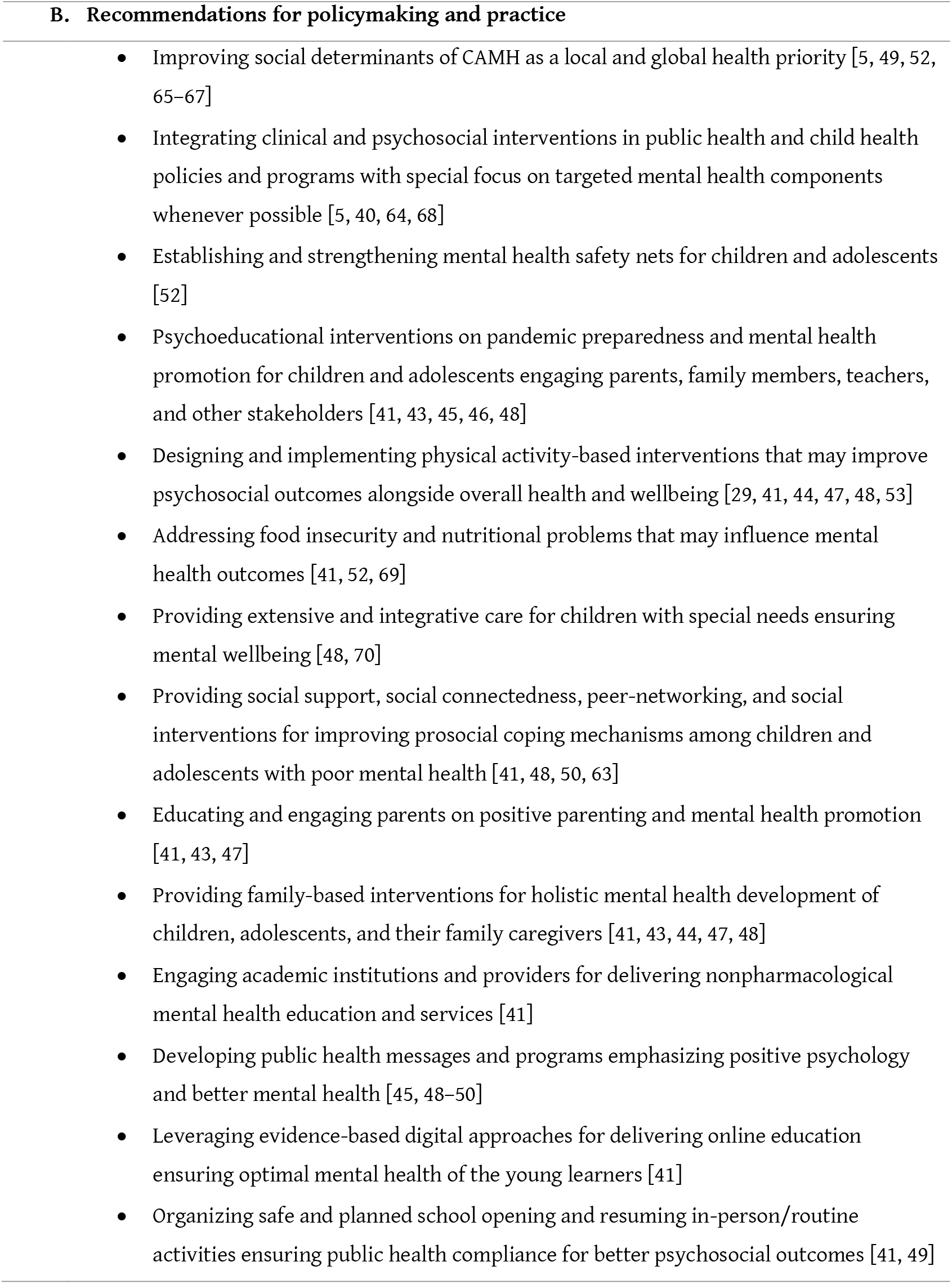
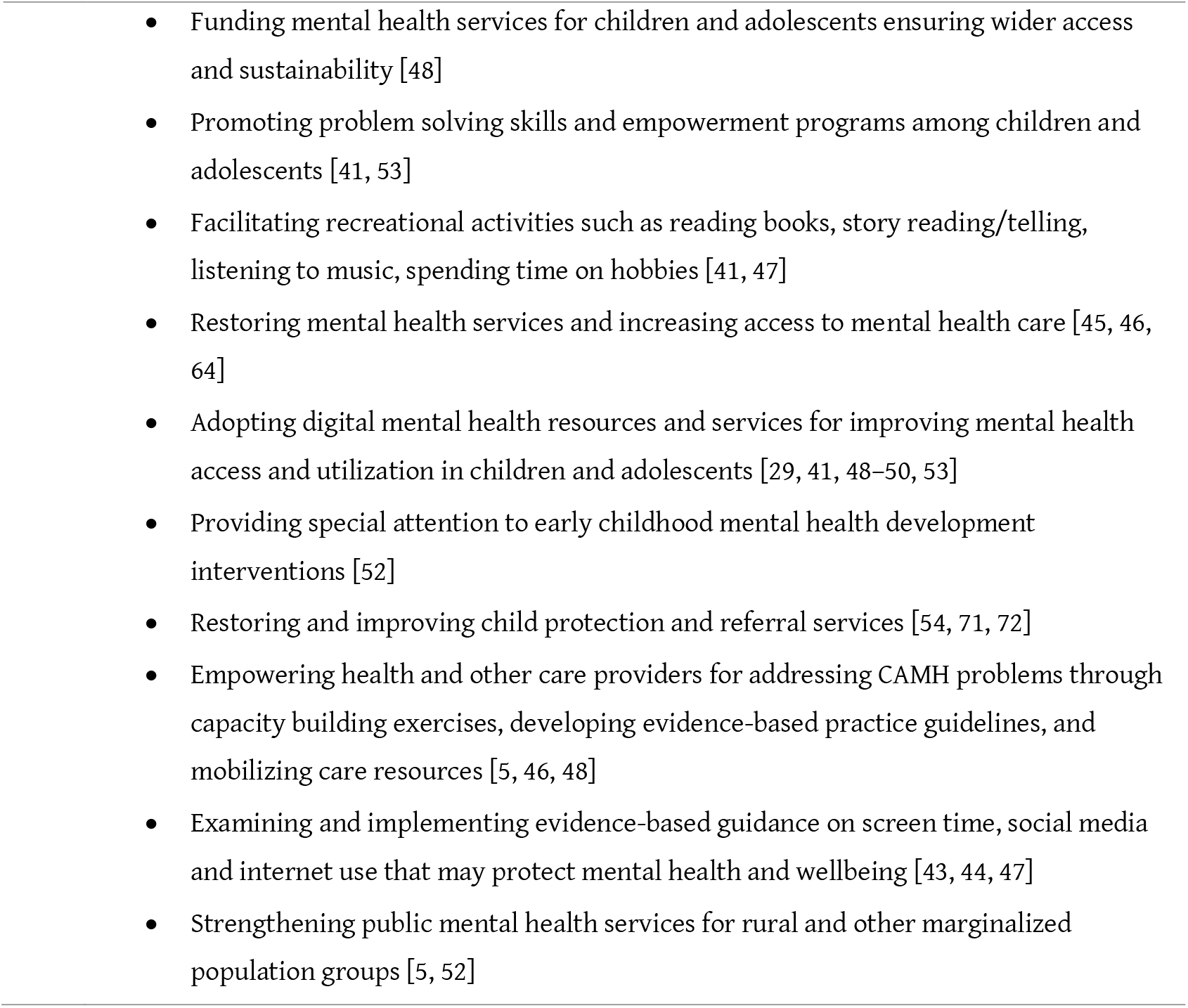
Evidence-based recommendations for future research, policymaking, and practice.

The current evidence on CAMH problems amidst COVID-19 necessitates multipronged efforts to alleviate the immediate and future health and social consequences of this burden. The policymakers and practitioners need to evaluate the evidence-based guidelines and recommendations that may fit the mental health needs of children and adolescents in different contexts, as listed in Table 3-B. At the individual level, awareness of mental health and pandemic safety should be strengthened, addressing the fear of infection and health hazards [41, 43, 45, 46]. Moreover, positive psychology-based targeted interventions should be promoted alongside public mental health interventions [40, 52, 64]. Furthermore, parents, teachers, and other caregivers should be engaged in mental health promotion [41, 43, 44, 47]. As many scholars suggest, those formal and informal caregivers may require psychoeducational and behavioral support to stay healthy and contribute to interpersonal and community health efforts [44, 47, 48]. Also, healthcare professionals and social workers should be empowered to offer psychosocial care services to the affected children and adolescents in diverse settings [46, 48]. Institutional care services should be made available and accessible to address the growing burden of mental health epidemics accompanying COVID-19 pandemic [40, 64]. Health systems and services must the strengthened to actualize these efforts at the societal level [45, 46, 64]. Lastly, a stronger policy commitment is needed to improve social determinants of mental health in children and adolescents that may enable them to overcome the current mental health problems and develop resilience to better psychosocial outcomes in the future [48, 49, 52].

### Limitations

This umbrella review has several limitations that must be addressed in future evidence syntheses. First, we included major databases but did not include preprint servers and other potential sources that may contain eligible reviews. Second, we identified multiple reviews with different methods and synthesized them narratively. A patient-level data synthesis from individual studies could have provided a more accurate and less overlapping view of the evidence landscape. Third, as systematic reviews and meta-analyses included primary studies published before conducting those reviews, a synthesis of the reviews is likely to miss insights from the most recent primary studies. Lastly, heterogeneity and publication biases were not assessed in this narrative umbrella study. Understanding those and addressing the same should be prioritized in future meta-research.

## Conclusions

Psychosocial stressors associated with the COVID-19 pandemic have resulted in a wide range of mental health problems in children and adolescents. The current evidence suggests a high burden of anxiety, depression, psychological distress, PTSD, sleep disorders, suicidal behavior, addiction disorders, and other psychosocial problems that require evidence-based measures to address the same. Also, many of these problems may persist and develop further mental health crises that should be addressed by adopting multifaceted psychiatric, psychological, and public health efforts. Transdisciplinary mental health research, policymaking, and practice should be emphasized to mitigate the mental health impacts of this pandemic and ensure optimal mental, physical, and social health of children and adolescents locally and globally.

## Data Availability

All data were retrieved from published literature that are available in the manuscript.

---

**Supplementary Material 1:**
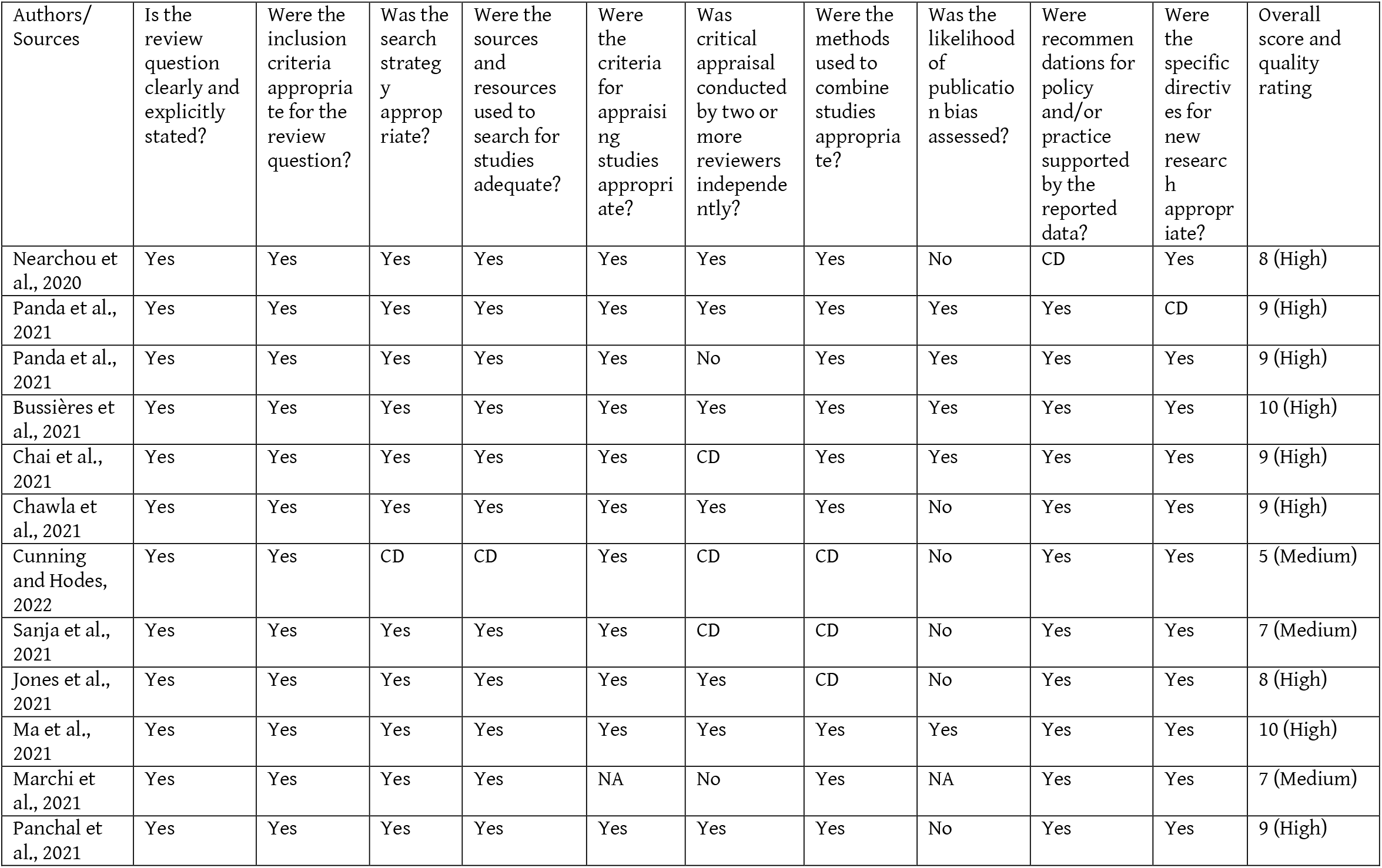
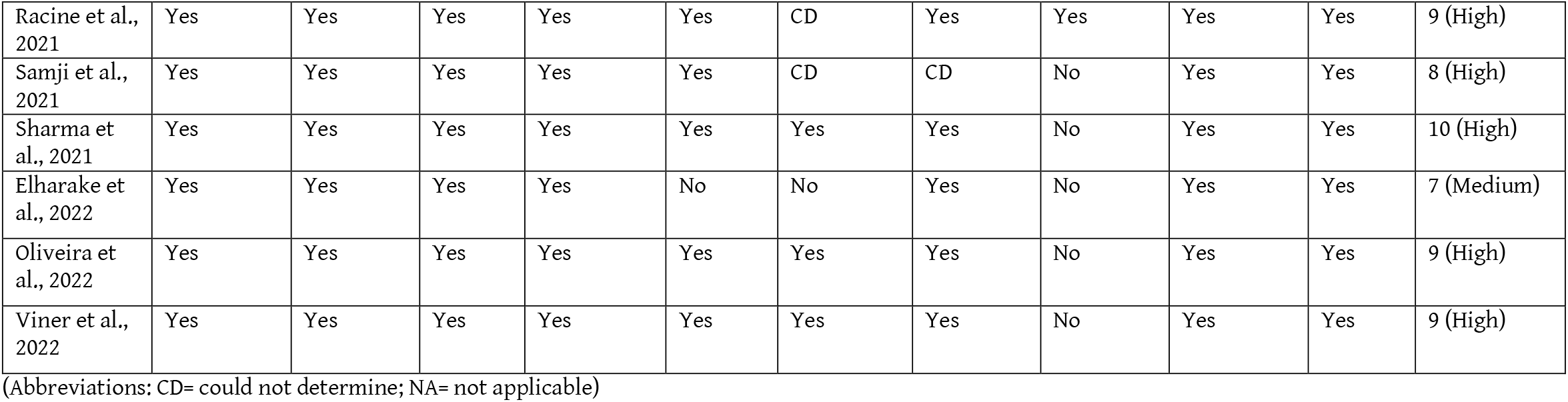
Critical appraisal of the methodological quality of the included reviews.

